# Similar costs and outcomes for differentiated service delivery models for HIV treatment in Uganda

**DOI:** 10.1101/2021.06.22.21259341

**Authors:** Teresa Guthrie, Charlotte Muheki, Sydney Rosen, Shiba Kanoowe, Stephen Lagony, Ross Greener, Jacqueline Miot, Hudson Balidawa, Josen Kiggundu, Jacqueline Calnan, Seyoum Dejene, Thembi Xulu, Ntombi Sigwebela, Lawrence C Long

**Author notes:** **Corresponding author:** Teresa Guthrie, Health Economics and Epidemiology Research Office, University of the Witwatersrand, 39 Empire Road, Johannesburg, South Africa. 2193.

## Abstract

**Background:** This study aimed to measure the total annual cost per patient and total cost per patient virally suppressed (defined as <1000 copies/ml) on antiretroviral therapy in Uganda in five differentiated service delivery models (DSDMs), including facility- and community-based models and the standard of care.

**Methods:** A cost/outcome study was undertaken from the perspective of the service provider, using retrospective patient record review of a cohort of patients over a two-year period, with bottom-up collection of patients’ resource utilization data, top-down collection of above-delivery level and delivery-level providers’ fixed operational costs, and local unit costs.

**Results:** Forty-seven DSDMs located at facilities or community-based points in four regions of Uganda were included in the study, with 653 adults on ART (>18 years old) enrolled in a DSDM. The study found that retention in care was 98% for the sample as a whole [96-100%], and viral suppression, 91% [86%-93%]. The mean cost to the provider (Ministry of Health or NGO implementers) was $152 per annum per patient treated, ranging from $141 to $166. Differences among the models’ costs were largely due to patients’ ARV regimens and proportions of patients on second line regimens. Service delivery costs, excluding ARVs, other medicines and laboratory tests, were modest, ranging from $9.66-16.43 per patient.

**Conclusion:** We conclude that differentiated ART service delivery in Uganda achieved excellent treatment outcomes at a cost similar to the standard of care. While large budgetary savings might not be immediately realized, the reallocation of “saved” staff time could improve health system efficiency as facilities and patients gain more experience with DSD models.

## Introduction

In 2019, there were an estimated 1.5 million adults living with HIV (PLHIV) in Uganda, equivalent to an HIV adult prevalence of 5.8% [5.4–6.2%] (UNAIDS 2020). Approximately 84% of the HIV-positive population were reported to be on antiretroviral therapy (ART), and of those, 75% were virally suppressed (UNAIDS 2020, Uganda AIDS Commission 2019). In Uganda, as in other high HIV prevalence countries, there is a need to adapt service delivery approaches to the needs and preferences of PLHIV, with the goal of maintaining good clinical outcomes, reducing costs to patients, and improving efficiency in service delivery (Ugandan Ministry of Health 2018a).

The Ugandan Ministry of Health (MOH) began piloting and scaling-up “differentiated ART service delivery models” (DSDMs) in 2016, becoming one of the first sub-Saharan African countries to develop and implement a comprehensive DSDM program. National guidelines for DSDMs were issued in 2017 (Ugandan Ministry of Health 2018a). As of 2018, there were five officially approved DSDMs in Uganda for both stable and complex ART patients (Box 1): facility-based individual management (FBIM), which is similar to the previous non-differentiated standard of care; facility-based groups (FBG); fast-track drug refills (FDR); community client-led ART delivery (CCLAD); and community drug distribution points (CDDP). By June 2020, roughly 79% of all adult ART patients had been enrolled in one of the five models: 42% in FDR, 34% in FBIM, 12% in FBGs, 7% in CCLAD, and 5% in CDDP (Kiggundu et al 2020). (The remaining 21% of patients were not recorded as being enrolled in a DSDM and are assumed to have been receiving standard of care treatment at facilities.) [Text Box 1].

There have been a few prior evaluations of the clinical outcomes of early versions of DSDMs in Uganda (Jaffar et al 2009; Weidle et al 2006; Ssuuna et al 2018; Kaimal et al 2017; Long et al 2020a), but there is little program-wide evidence on costs and effectiveness, a dearth that limits national budgeting, resource mobilization, implementation planning, and scale-up. At the request of the Ugandan MOH, the PEPFAR-funded EQUIP Project conducted a cost-outcome analysis of the five DSDMs to estimate the annual cost per person retained in care and per patient virally suppressed in each model.

### Box 1

**Differentiated ART service delivery models in Uganda**

i. Facility-based individual management (FBIM): for patients needing extra attention, such as unstable/complex patients, those who have recently been initiated in care, and those who chose to continue to receive their services at the facility. FBIM is the conventional standard-of-care model of ART delivery.
ii. Facility-based groups (FBG): for stable or complex patients needing peer support, such as adolescents, pregnant and breastfeeding women (PBFW), and discordant couples. The frequency of their ARV refills depends on patients’ stability.
iii. Fast-track drug refills (FDR): for stable patients who pick-up their ARVs directly from clinics (and these can include patients on second-line regimens).
iv. Community client-led ART delivery (CCLAD): stable patients form groups within their communities. One person is selected (on rotational basis) to collect the ARV refills for the whole group from the facility.
v. Community drug distribution points (CDDP): stable patients pick up their ARVs from a community outreach point, including private pharmacies (Kiggundu et al 2020).

Longer appointment spacing and multi-month scripting can be offered to stable patients in all models.

## Methods

In this study, we estimated the annual cost per patient outcome of a cohort of Ugandan ART patients enrolled in the five official DSDMs in 2017. The cost to providers for individual patient resource use was estimated using a bottom up, micro costing approach, with retrospective data drawn from patients’ medical records using methods previously described (Rosen et al 2008; Long et al 2011; Long et al 2017). Public and non-public (private not-for-profit) providers’ and implementing partners’ fixed and shared operational, management, and supervisory costs were estimated using a top-down approach. We refer to two periods of observation (study periods): 0-12 months after study enrolment, which corresponds to calendar year 2017 (1 January 2017-31 December 2017), and 13-24 months after study enrolment, which corresponds to calendar year 2018 (1 January 2018-31 December 2018). These are study observation periods only; they do not refer to patients’ duration on ART or time in the DSDM. Costs are reported from the provider’s perspective only.

### Study sites

Study sites were selected to capture the variation in settings, implementing partners, and other characteristics of ART services in Uganda. We define a “site-model” as one model being implemented by one ART facility, though model services may be delivered at non-facility locations. Using this definition, a facility can have more than one site-model if more than one differentiated model is offered there. Our sampling frame included any site-model which had been in operation for ≥6 months by January 2017. Site-models that were considered outliers in terms of size (number of patients) or access (extreme locations that were physically difficult to reach) were excluded from the sampling frame.

In January 2017, there were 605 site-models that met our sample criteria at 297 facilities in Uganda. From these, multi-stage purposive sampling was used to select 47 site-models so as to reflect variation according to model type, facility ownership (public and private-not-for-profit), patient volume, geographic location, and implementing partner (further details on the sampling criteria are included in Supporting information S1 File). We note that many of the public facilities in the study were supported by a non-governmental “implementing partner” receiving external donor support largely from PEPFAR. These implementers played a major role in establishing and maintaining the DSDMs. We thus captured their operational costs, as well as those of the MOH.

Fixed costs for providers and implementers were collected for all 47 site-models. Twenty of the 47 (4 per DSDM) were then selected for the collection of patient level resource usage and treatment outcomes.

### Study population

Our study population included all adult ART patients (≥18 years) who were enrolled in a DSDM on or before 1 January 2017. In Uganda, all PLHIV are eligible for DSDMs, but their specific model options depend on model availability and clinical stability. A “stable” patient is defined as one who is a) on their current ART regimen for ≥12 months; b) virally suppressed; c) in WHO Stage I/II; d) adherent (> 95%) over the last 6 months; and e) if a TB patient, past the intensive TB treatment phase (2 months) and sputum negative (Kiggundu 2020). (We note that Uganda refers to ART patients as “clients,” but we have chosen to use “patients” here for consistency with the international literature.) Patients who met these eligibility criteria were selected consecutively from DSDM registers kept by the facilities starting in January 2017 and then sequentially earlier in time (December 2016, November 2016, and so on) until the target sample size of 30-33 patients was reached for each of the 20 sites. Patients with a record of formal transfer out of a selected health facility before the 12-month study endpoint were excluded. For the FBG sites, only groups for pregnant and breastfeeding women (PBFW) were selected because of more rigorous ethical clearance requirements for accessing pediatric and youth groups and the small number of sero-discordant couple groups.

Participants in each of the models except FBGs were followed longitudinally for 24 months starting on January 1, 2017. This follow-up period was broken into two periods: 0-12 months after study enrolment and 13-24 months after study enrolment, with data accessed retrospectively at the end of each period. For the FBGs, two different samples of PBFW were followed for each 12 month period (FBG1 and FBG2) because they only remained in their FBG for the duration of their pregnancy and postnatal period.

### Data collection

All data for the study were collected locally from three sources. First, research assistants retrospectively extracted demographic characteristics, medical histories, treatment outcomes, dates of and reasons for clinic visits, ARVs and non-ARV medications dispensed, TB status, WHO clinical stage, laboratory tests, and counselling sessions from study participants’ ART care cards, which were maintained by facility staff. Second, from model-specific DSDM registers, participants’ attendance at any DSDM-related event were recorded (adherence counselling, group support meetings, FBG meetings, community medication collection/distribution meetings, community viral load sessions etc). Third, we interviewed programme and financial managers at each of the site-models’, collected the estimated length of time spent by the different cadre for each service, obtained expenditure records, asset registers and undertook spacial measurements of the buildings used in providing the DSDM services.

### Treatment outcome measures

Retention in care and viral suppression as reported in individual participants’ medical records were the primary treatment outcomes of interest in this analysis. Retention was measured as not having missed a scheduled appointment (clinic or DSDM) for >90 consecutive days (Ugandan Ministry of Health 2018a). Viral suppression was based on the latest viral load (VL) test in each study period (12/24 month ± 3-month window) being <1000 copies/ml (Ugandan Ministry of Health, 2018).

For the cost analysis, we defined four mutually exclusive outcomes as follows: Retained in care and known to be virally suppressed (RIC, suppressed); retained in care and known to be not virally suppressed (RIC, unsuppressed); retained in care and VL unknown (RIC, VL unknown); not in care (NIC). ARV adherence, as proxied by an annual medication possession ratio (MPR) (total days dispensed/365), was reported as a secondary treatment outcome and categorized using the MOH’s scale (good ≥95%; fair 85%-94%; poor 75%-84%; non-adherent ≤74%). Patients in the cohort who switched between models during the study period were retained in their original models for analysis.

### Resource utilization, cost data and cost analysis

To calculate direct resource utilization for each patient, we identified and quantified all resources utilized within the two 12-month study periods. Patient-level resource utilization data were identified and quantified from patients’ ART care cards and DSDM registers, as described above. The cost per unit of each resource were collected from price lists, salary scales, tender documents, and implementers’ expenditure logs. Staff costs per facility visit or DSDM event were calculated based on the estimated time per visit or DSDM event for each staff member at the average cost of that cadre’s time, based on total remuneration. The estimated time per visit was estimated from staff interviews. Quantities of resources used were multiplied by unit costs and summed to obtain an average direct cost per patient. (Details of prices and costing methods are described in Supporting information S1 Table and S2 Table).

We also estimated indirect (fixed and shared) costs, including facility and DSDM management, administration, oversight and supervision, staff training, equipment, building/ rental and all operational and overhead costs at the facility and above-facility levels. These indirect costs, varying by model type, were attributed to each DSDM patient using an allocation factor based on facility annual headcount (out-patient visits) and each patient’s number of visits.

Finally, we summed the direct and indirect cost/patient to generate a total cost per patient, stratified by DSDM-type and patient outcome. We also estimated the “production cost” of achieving one patient who was virally suppressed by dividing the total cost (any outcome) per model by the proportion of patients with viral suppression in that model.

Unit costs reflect 2018 market prices and were converted from Uganda shillings to United States dollars (USD) using the annual average Bank of Uganda exchange rate for 2018 of $1:UGX 3728 (Bank of Uganda 2019). Costs are reported in 2018 USD.

## Results

### Study population

A total of 653 patients from four regions of Uganda were enrolled in the study, divided roughly evenly among the five DSDM types (Table 1). During the two-year study period, 29 patients switched back to FBIM due to viral failure, while 6 FBIM patients switched to other DSDMs. As explained above, these patients were retained in their original models for purposes of analysis. The majority (473, 72%) were female, a slightly higher proportion than in the national ART cohort (65%) (Ugandan Ministry of Health 2018b), due to our sampled FBG participants being all female. The facility-based individual models (FDR and FBIM) had the highest proportions of male participants: 58 (44%) and 46 (36%), respectively. The median age for all the models except FBG ranged from 41 to 44 years; FBG patients were younger, with a median age of 29 years. The median duration on ART was 5 years; FBIM and FBG patients had been on ART for less time (2 and 3 years respectively), while FDR patients had spent a median of 8 years on ART. At study enrolment, the median length of time in a differentiated model was one year, and 593 (91%) of patients were on first line (FL) ARV regimens. Only the FDR model cohort reported more than 10% of participants on a second line (SL) regimen (22, 17%). (Table 1).

**Table 1:** Characteristics and treatment outcomes by model of ART delivery.

| Sample characteristics (n, % unless otherwise specified) | FBIM<br>(n=128) | CCLAD<br>(n=131) | CDDP<br>(n=132) | FBG1 /2<br>(n=129, 115)<sup>a</sup> | FDR<br>(n=133) | Total<br>(n=653) |
| --- | --- | --- | --- | --- | --- | --- |
| Sex (female) | 82 (64%) | 92 (70%) | 95 (72%) | 129 (100%) | 75 (56%) | 473 (72%) |
| Age, years (median, IQR)* | 41 (34-51) | 44 (40-49) | 44 (38-52) | 29 (25-34) | 44 (35-51) | 41 (33-48) |
| Duration on ART, years (median, IQR)* | 3 (2-5) | 5 (2-8) | 7 (5-10) | 2 (1-3) | 8 (5-10) | 5 (2-8) |
| Duration in DSDM, years (median, IQR)<sup>b</sup> | 3 (2-5)<sup>c</sup> | 1 (1-1) | 1 (1-6) | 1 (1-1) | 2 (1-4) | 1 (1-3) |
| Patients on first-line regimens<sup>b</sup> | 117 (91%) | 124 (95%) | 121 (92%) | 120 (93%) | 111 (83%) | 593 (91%) |
| Patients on second-line regimens<sup>b</sup> | 11 (9%) | 7 (5%) | 11 (8%) | 9 (7%) | 22 (17%) | 60 (9%) |
| Baseline CD4 count, cells/μl (median, IQR)<sup>d</sup> | 310<br>(199-430) | 221<br>(128-353) | 210<br>(143-328) | 433<br>(250-629) | 234<br>(118-349) | 272<br>(152-414) |
| **Outcomes at 12 months** |  |  |  |  |  |  |
| Retained in care | 126 (98%) | 127 (97%) | 130 (98%) | 120 (93%) | 133 (100%) | 636 (97%) |
| Unsuppressed (viral load >1000 copies/ml) | 12 (9.4%) | 4 (3.1%) | 3 (2.3%) | 6 (4.7%) | 4 (3%) | 29 (4.5%) |
| Suppressed (viral load <1000 copies/ml) | 110 (86%) | 125 (95%) | 123 (93%) | 115 (89%) | 118 (89%) | 591 (91%) |
| Unknown viral status | 6 (5%) | 2 (2%) | 1 (1%) | 8 (6%) | 11 (9%) | 28 (4.3%) |
| **Outcomes at 24 months** |  |  |  |  |  |  |
| Retained in care<sup>e</sup> | 122 (97%) | 127 (98%) | 132 (100%) | 110 (96%) | 131 (99%) | 622 (98%) |
| Detectable viral load (>1000 copies/ml) | 10 (7.9%) | 2 (1.5%) | 3 (2.3%) | 4 (3.5%) | 3 (3%) | 22 (3.5%) |
| Undetectable viral load (<1000 copies/ml) | 111 (88%) | 117 (90%) | 121 (92%) | 108 (94%) | 119 (90%) | 576 (91%) |
| Unknown viral status | 5 (4%) | 11 (9%) | 8 (7%) | 3 (3%) | 10 (8%) | 37 (6%) |

<sup>a</sup> Sample characteristics and 12-month outcomes are for the FBG1 cohort; 24 month outcomes are for the FBG2 cohort.
<sup>b</sup> Age, duration on ART, duration on DSDM and regimen are measured at the time of enrollment in the study (January 2017).
<sup>c</sup> For FBIM, the duration on DSDM is equivalent to the duration on ART. Some patients switched from their DSDM back to FBIM when becoming unsuppressed, but they were retained in their original models for purposes of analysis.
<sup>d</sup> Baseline CD4 count at time of ART initiation. Data missing for <10% of patients in all models.
<sup>e</sup> Not retained in care (in 2<sup>nd</sup> year period) included one death (FBIM); the rest were LTFU.

### Treatment outcomes

Overall retention in care and viral suppression rates were high for all the models (Table 1). For the sample as a whole, retention in care was 97% and 98% at 12 and 24 months, respectively; average viral suppression was 91% for both periods. FBIM patients had the highest proportion of known non-suppressed patients (9.4% and 7.9%) and was the only model to report a death, which occurred in the second study period, while FBG had the highest suppression rate at 94%. The majority of patients in both study periods (80% and 83%, respectively) were classified as having “good” adherence (≥95%), based on the annual MPR and the scale provided by the MOH (where the FBIM mean ARV days prescribed for the year were 364, CCLAD 361, CDDP 363, FBG 364 and FDR 369).

### Resource utilization

#### Antiretroviral medications and laboratory tests

A range of ARV formulations were prescribed and dispensed to our study participants. The most common at 24 months were TDF-3TC-EFV for first line therapy, which accounted for 50% of first line formulations, and TDF-3TC-ATV/r for second line therapy, accounting for 27% of second line regimens (Supporting information S3 Table). Dolutegravir (DTG) became available in 2018; 8.2% of patients had switched to DTG formulations by the 24^th^ study month. Patients received an average of 1-2 months of ARVs at a time—there was little adoption of multi-month dispensing during the study periods. The annual MPR was high, with some clients receiving more than 365 days of ARVs over the year.

Viral load testing appeared consistent with guidelines: study participants received an average of one viral load test per year, and with only minor variation by model (Table 2). There was a reduction in other laboratory investigations between the study periods, from 0.62 tests per patient in 0-12 months to 0.28 tests per patient in 13-24 months (refer to Supporting information S4 Table).

**Table 2:**
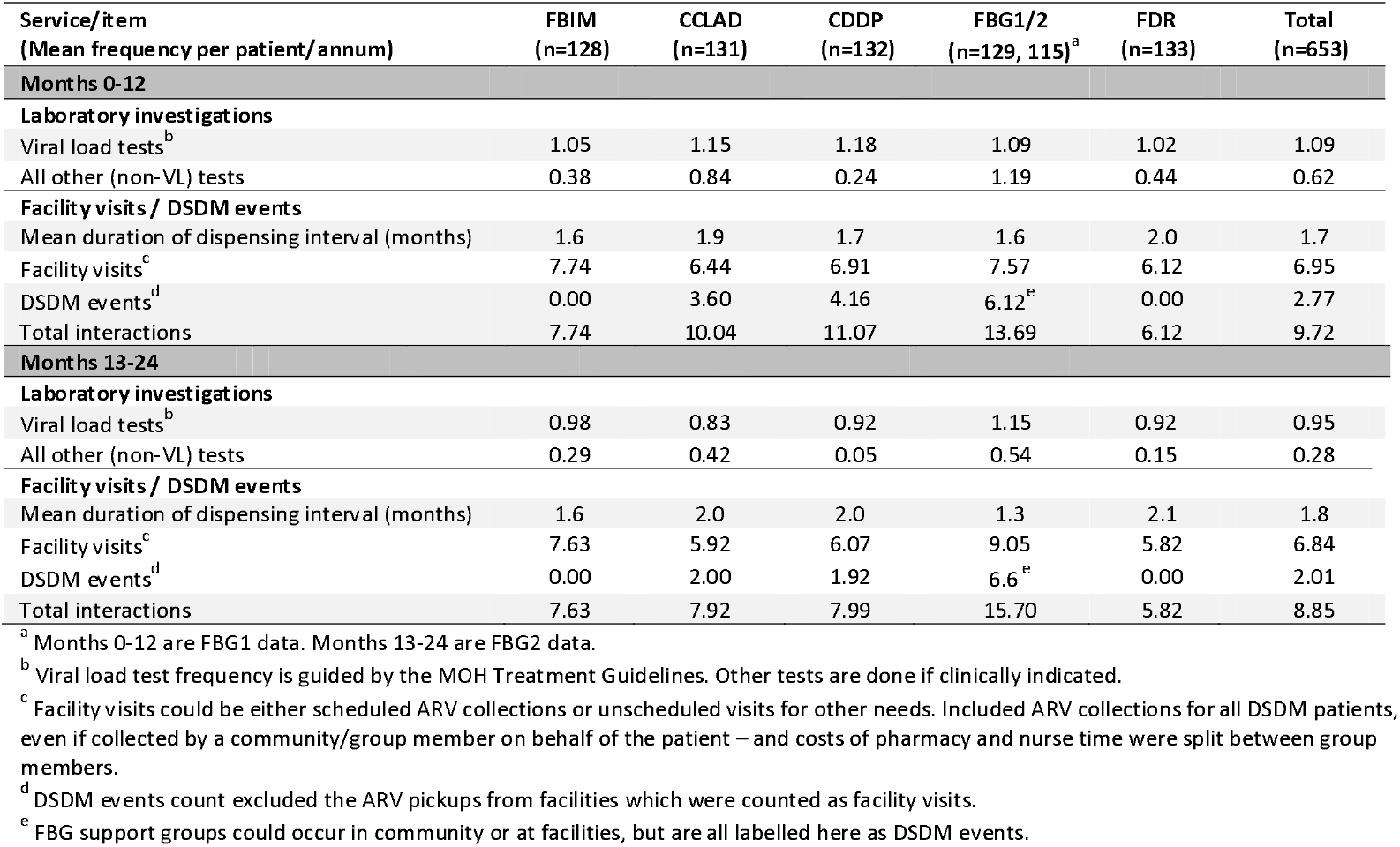
Resource utilization per patient, by model and study period (mean per patient per annum)

#### Frequency of facility visits and DSDM events

Patients visited healthcare facilities during the study period either for a scheduled (routine) appointment to collect their ARVs (individual collection, group collection, or fast-track drug refill) or for unscheduled visits for HIV-related illnesses, opportunistic illnesses, or other comorbidities (Table 2). In addition to facility visits, the CCLAD, CDDP, and FBG models held DSDM-specific events, or interactions, such as community-based clinical/TB assessments, group viral load sessions, ARV collections, and adherence support meetings (Supporting information S5-S6 Tables). The available data indicated a reduction of almost half (48%) in the total recorded DSDM events between the two study periods, which may reflect actual changes in patients’ participation, deterioration in record keeping, or both. Actual implementation of the DSDM models differed slightly from MOH guidelines (Kiggundu 2020) in the frequency of facility ART visits, DSDM interactions, and viral load tests.

These differences diminished over the two-year study period, as greater standardization occurred in DSDM implementation.

### Total cost per patient and cost per outcome

Unit costs of the resources utilized by participating patients are available in Supporting information S1, S3-S6 Tables). For the second study period, which may better reflect costs going forward, the annual mean cost per patient treated was $141, $146, $150, $152 and $166 for the FBG, CDDP, CCLAD, FBIM (standard of care) and FDR models, respectively (Table 3). FBIM and FDR costs were largely driven by having greater proportions of patients on second-line regimens (9% and 17% respectively, by the end of the study period). The mean annual cost per second-line (SL) patient across all models was more than double that of first-line (FL) patient ($135 FL vs. $343 SL). The mean cost per virally suppressed patient (at 24 months after study enrolment) was $150, $158, $167, $173 and $184, for FBG, CDDP, CCLAD, FBIM and FDR respectively.

**Table 3.**
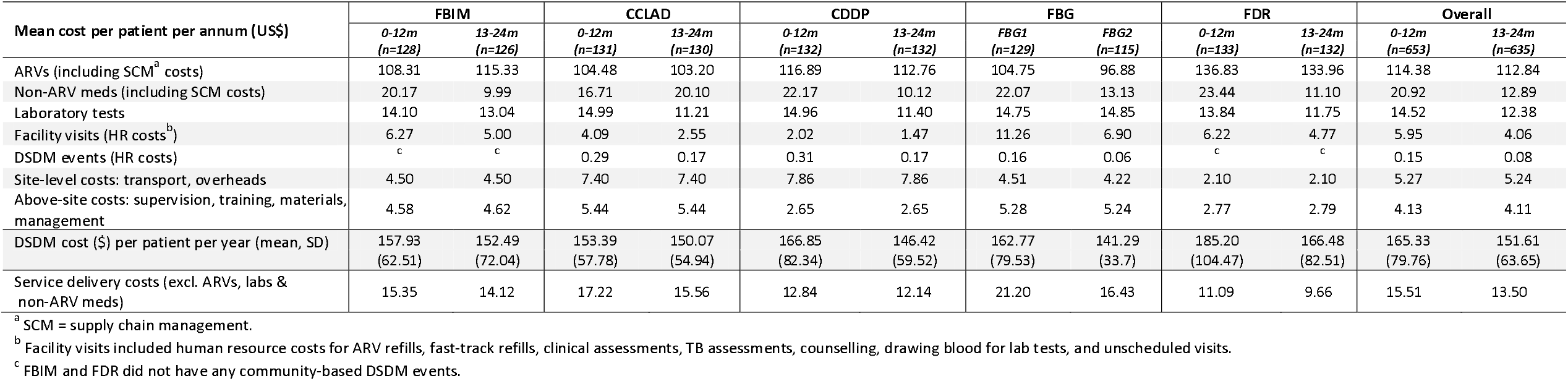
Annual average cost per patient by cost component, by model and period (US$ 2018)

| Mean cost per patient per annum (US\$) | FBIM | | CCLAD | | CDDP | | FBG | | FDR | | Overall | |
| --- | --- | --- | --- | --- | --- | --- | --- | --- | --- | --- | --- | --- |
|  | 0-12m<br>(n=128) | 13-24m<br>(n=126) | 0-12m<br>(n=131) | 13-24m<br>(n=130) | 0-12m<br>(n=132) | 13-24m<br>(n=132) | FBG1<br>(n=129) | FBG2<br>(n=115) | 0-12m<br>(n=133) | 13-24m<br>(n=132) | 0-12m<br>(n=653) | 13-24m<br>(n=635) |
| ARVs (including SCM<sup>a</sup> costs) | 108.31 | 115.33 | 104.48 | 103.20 | 116.89 | 112.76 | 104.75 | 96.88 | 136.83 | 133.96 | 114.38 | 112.84 |
| Non-ARV meds (including SCM costs) | 20.17 | 9.99 | 16.71 | 20.10 | 22.17 | 10.12 | 22.07 | 13.13 | 23.44 | 11.10 | 20.92 | 12.89 |
| Laboratory tests | 14.10 | 13.04 | 14.99 | 11.21 | 14.96 | 11.40 | 14.75 | 14.85 | 13.84 | 11.75 | 14.52 | 12.38 |
| Facility visits (HR costs<sup>b</sup>) | 6.27 | 5.00 | 4.09 | 2.55 | 2.02 | 1.47 | 11.26 | 6.90 | 6.22 | 4.77 | 5.95 | 4.06 |
| DSDM events (HR costs) | <sup>c</sup> | <sup>c</sup> | 0.29 | 0.17 | 0.31 | 0.17 | 0.16 | 0.06 | <sup>c</sup> | <sup>c</sup> | 0.15 | 0.08 |
| Site-level costs: transport, overheads | 4.50 | 4.50 | 7.40 | 7.40 | 7.86 | 7.86 | 4.51 | 4.22 | 2.10 | 2.10 | 5.27 | 5.24 |
| Above-site costs: supervision, training, materials, management | 4.58 | 4.62 | 5.44 | 5.44 | 2.65 | 2.65 | 5.28 | 5.24 | 2.77 | 2.79 | 4.13 | 4.11 |
| DSDM cost (\$) per patient per year (mean, SD) | 157.93<br>(62.51) | 152.49<br>(72.04) | 153.39<br>(57.78) | 150.07<br>(54.94) | 166.85<br>(82.34) | 146.42<br>(59.52) | 162.77<br>(79.53) | 141.29<br>(33.7) | 185.20<br>(104.47) | 166.48<br>(82.51) | 165.33<br>(79.76) | 151.61<br>(63.65) |
| Service delivery costs (excl. ARVs, labs & non-ARV meds) | 15.35 | 14.12 | 17.22 | 15.56 | 12.84 | 12.14 | 21.20 | 16.43 | 11.09 | 9.66 | 15.51 | 13.50 |

<sup>a</sup> SCM = supply chain management.
<sup>b</sup> Facility visits included human resource costs for ARV refills, fast-track refills, clinical assessments, TB assessments, counselling, drawing blood for lab tests, and unscheduled visits.
<sup>c</sup> FBIM and FDR did not have any community-based DSDM events.

ARVs and laboratory tests were the main cost drivers for all models – 74% and 9% respectively of total costs (Table 3) - followed by the prevention and treatment of opportunistic infections which included Isoniazid and Pyridoxine (8% on average). If these three cost components are removed from the totals, the mean annual service delivery cost per patient was $10 for FDR, $12 CDDP, $14 FBIM, $16 FBG, and $16 for CCLAD. Human resource costs for facility visits (3% on average) varied across the models, based on the different staff involved, their salary scales, and the length of time and frequency of each interaction. Participants in the FBGs (pregnant and breastfeeding women) appeared to have a greater proportion of personnel costs, due to more frequent facility visits and interactions. Human resource costs for the DSDM events/interactions were low (0.1%) because most were group events for which staff costs were shared among the group participants. Site overhead costs (3%) and above-site costs (3%), for supervision, training, management, and implementing partners’ headquarter costs, varied between models but generally account for only a small share of the total per patient. CCLAD had slightly higher above-site costs than the other models, in part due to their greater supervision, monitoring and headquarters’ operating costs, while CDDP and FDRs had the lowest above-site costs (Table 3).

## Discussion

By 2018, Uganda had developed and implemented five differentiated models of ART service delivery, including the standard of care, known as facility-based individual management (FBIM). In this two-year observational study, we found that, on average, all five DSDMs achieved good outcomes and cost the provider (Ministry of Health or NGO implementers) an average of $152 per year per patient treated. Retention in care averaged a surprisingly high 98% for the sample as a whole, with a tight range of 96-100%. Viral suppression, which averaged 91%, varied between a low of 88% among patients in FBIM, which served as the primary model for treating complex patients, and a high of 94% among FBG patients.

These good outcomes are consistent with other reports on the CDDP (Ssuuna 2018; Kaimal 2017) and CCLAD (Weidle 2006) models in Uganda. Similarly, in other African countries, a recent systematic review concluded that retention in care and viral suppression are roughly equivalent to those in conventional models of care (Long et al 2020b).

Differences among the models’ costs were explained largely by patients’ ARV regimens and the costs of prevention and treatment of opportunistic infections and other co-morbidities. Service delivery costs, excluding ARVs, laboratory tests and other non-ARV medicines, were modest, ranging from $10-$16 per patient, with CCLADs being slightly higher due higher above-site costs while FBGs personnel costs were higher due to increased facility visits and interactions. This does not leave a lot of room for “savings” to the healthcare system, and indeed, we found the new Ugandan DSDMs were not much less expensive than FBIM, the model that most closely proxies the previous standard of care.

This finding is similar to results of some other recent studies but not with others. A recent observational evaluation in Zambia, for example, found that the standard of care model was less expensive than community-based ART delivery (Nichols et al 2020a). In South Africa, in contrast, a study of adherence clubs where lower cadre staff (compared to the facility-based standard of care staff) dispensed ARVs to 25-30 members at club meetings found them cost-saving compared with the standard of care (Bango et al 2016). Evaluations of models implemented in cluster-randomized trials that explicitly emphasized multi-month dispensing of ARVs have also observed modest cost savings (Nichols et al 2020b). We note that in Uganda, over the period of this study, participants made more facility visits for medication collection than called for in guidelines. Since the study ended, Uganda has implemented longer dispensing intervals for ARVs, which may lead to lower costs for models that are able to dispense six-month supplies to a large share of patients.

Numerous studies have also found that DSDMs do substantially reduce costs to patients, primarily for transport and time (Long 2020a). With equivalent or better outcomes and large benefits to patients, the finding that differentiated models do not greatly reduce provider costs may not diminish their societal value.

Our study had a number of limitations, largely stemming from our reliance on routinely-collected, retrospective data. Because of incomplete electronic patient medical records at some sites, we relied on individual patients’ paper ART Care Cards, which are removed from healthcare facilities when patients die. As a result, we likely undercounted deaths in the 0-12 month sample, and during the second study period, we identified only one death. Our outcomes measures were therefore limited to patients surviving at 24 months, possibly causing us to overstate rates of retention and viral suppression. We also struggled with incomplete records of DSDM interactions, as model registers were poorly maintained and this worsened in the second study period. The decrease in DSDM interactions, from an annual average 2.85 in the first period to 2.05 in the second period, may thus reflect either an actual reduction in DSDM interactions or a worsening in record-keeping between the years. Finally, estimates of staff time spent for each type of event were obtained through interviews with staff. Self-reported time use may not be accurate, and we excluded non-patient-facing activities such as record keeping, stock management, and breaks. We thus may have underestimated these human resource costs for every model. In a separate facility-level analysis of total salary costs/patient, we estimated an additional personnel cost of $2.20 per patient per year, for these non-patient-facing activities. These could be added to the totals for each model in Table 3.

In conclusion, differentiated ART service delivery in Uganda achieved excellent treatment outcomes at a cost similar to standard of care (FBIM). While large budgetary savings might not be immediately realized, the reallocation of “saved” staff time due to multi-month dispensing and reduced facility visits could improve health system efficiency as facilities and patients gain more experience with the DSD models.

## Data Availability

The data are owned by the study sites and Ministry of Health (Uganda) and governed by the TASO Research Ethics Committee (Uganda). All relevant data are included in the manuscript and supporting information files. The full data are available from HealthNet Consult (Uganda) for researchers who meet the criteria for access to confidential data and have approval from the owners of the data. Contact the organization representation Ms. Charlotte Muheki for additional information regarding data access.

## Abbreviations

CCLAD: community client-led ART delivery
CDDP: community drug distribution points FBG facility-based groups
FBIM: facility-based individual management
FDR: fast-track drug refills
IAC: intensive adherence counselling
LTFU: lost to follow-up
MMS: multi-month scripts
MPR: medication possession rate
PLW: pregnant and lactating women
VL: viral load
VS: virally suppressed

## References

Bango F, Ashmore J, Wilkinson L, van Cutsem G, Cleary S. 2016. Adherence clubs for long-term provision of antiretroviral therapy: cost-effectiveness and access analysis from Khayelitsha, South Africa. Trop Med Int Health. 21(9): 1115–23.

Bank of Uganda. 2019. Historical currency exchange rates. Available: https://www.bou.or.ug/bou/bouwebsite/BOU-HOME [accessed 10 January, 2019].

Jaffar S, Amuron B, Foster S, et al. 2009. Rates of virological failure in patients treated in a home-based versus a facility-based HIV-care model in Jinja, southeast Uganda: a cluster-randomised equivalence trial. The Lancet. 374(9707): 2080–9.

Kaimal A, Castelnuovo B, Atwiine M, Musomba R, Nabaggala MS, Ratanshi RP, Lamorde M. 2017. Experiences with Retention in Care and Viral Suppression in a Pharmacy Refill Program. Presentation at Conference on Retroviruses and Opportunistic Infections.

Kiggundu J, Katureebe C, Balidawa H, Lukabwe I. 2020. DSD updates in Uganda: Uganda Ministry of Health AIDS Control Program. Presented 29 Sept 2020 to the DSD Technical Working Group.

Long L, Brennan A, Fox MP, Ndibongo B, Jaffray I, Sanne I, et al. 2011. Treatment outcomes and cost-effectiveness of shifting management of stable ART patients to nurses in South Africa: an observational cohort. PLoS Med. 8(7):e1001055. [PubMed: 21811402].

Long LC, Maskew M, Brennan AT, et al. 2017. Initiating antiretroviral therapy for HIV at a patient’s first clinic visit: a cost-effectiveness analysis of the rapid initiation of treatment randomized controlled trial. AIDS. 31(11):1611–1619. doi:10.1097/QAD.0000000000001528.

Long L, Kuchukhidze S, Pascoe S, et al. 2020a. Differentiated service delivery models for antiretroviral treatment of HIV in sub-Saharan Africa: A rapid systematic review. Boston: Boston University and HERO.

Long L, Kuchukhidze S, Pascoe S, Nichols BE, Cele R, Govathson C, Huber A, Flynn D, Rosen S. 2020b. Retention in care and viral suppression in differentiated service delivery models for HIV treatment in sub-Saharan Africa: a rapid systematic review. J Int AIDS Soc. Forthcoming.

Nichols BE, Cele R, Jamieson L, Long L, Siwale Z, Banda P, Moyo C, Rosen, S. 2020a. Community-based delivery of HIV treatment in Zambia: costs and outcomes (pending publication).

Nichols B, Fatti G, Cele R, Lekodeba N, Maotoe T, Sejana MV, Chasela C, Faturiyele I, Tukei B, Rosen S. 2020b. Economic evaluation of differentiated service delivery models for ART service delivery from a cluster-randomized trial in Lesotho: Cost to provider and cost to patient. Abstract PEE1626, AIDS 2020, July 6-10. Presentation of INTERVAL results at CQUIN webinar.

Rosen S, Long L, Sanne I. 2008. The outcomes and outpatient costs of different models of antiretroviral treatment delivery in South Africa. Trop Med Int Health. 13(8):1005–15. [PubMed: 18631314].

Ssuuna M, Nakade S, Zalwango S, Mubiru J, Okello D, Otim L, Kigozi J. 2018. The IDI-KCCA Community Pharmacy ART Refill Program. Poster Presentation.

Ugandan AIDS Commission. 2019. Presidential Fast-Track Initiative on Ending HIV and AIDS in Uganda: Factsheet. Available from: www.uac.go.ug

Ugandan Ministry of Health. 2017. Implementation Guide for Differentiated Service Delivery of HIV Services in Uganda.

Ugandan Ministry of Health. 2018a. Consolidated Guidelines for Prevention and Treatment of HIV in Uganda.

Ugandan Ministry of Health. 2018b. HIV Care and Treatment summaries for October – December, 2018.

UNAIDS. 2020. AIDS 2020 Data Book. Available from: https://www.unaids.org/sites/default/files/media_asset/2020_aids-data-book_en.pdf.

Weidle PJ, Wamai N, Solberg P, et al. 2006. Adherence to antiretroviral therapy in a home-based AIDS care programme in rural Uganda. The Lancet. 368(9547): 1587–94.

