## Supporting information for "Similar costs and outcomes for differentiated service delivery models for HIV treatment in Uganda" (from another version of this preprint: version 1, 2021-06-28)

**S1 Table. Unit costs for human resources for services, laboratory tests and ARV formulations**

| **Service / event / item** | | | **Mean cost**  **(US$ 2018)** |
| --- | --- | --- | --- |
| **Human resource costs for facility-based services** | | | **Per service** |
| Drug refill | | | 0.09 |
| Fast-track drug refill | | | 0.19 |
| Comprehensive clinical assessment | | | 1.21 |
| TB assessment at clinic | | | 1.52 |
| Unscheduled clinic visit for OIs, co-morbidities, other | | | 1.71 |
| Drawing blood at clinic | | | 0.23 |
| FBG quick clinical assessment | | | 0.09 |
| Counselling: education on basic HIV prevention and disclosure | | | 1.52 |
| Counselling: ART preparation, initiation, support, monitoring | | | 1.40 |
| Counselling: Progression on treatment | | | 1.34 |
| Intensive adherence counselling | | | 2.60 |
| **Human resource costs for community-based services** | | | **Per service** |
| CCLAD pre-med/meds collection/support meetings* | | | - * |
| Viral Load testing meeting (CCLAD & CDDP)** | | | 0.11 |
| Counselling session (CCLAD & CDDP)** | | | 0.04 |
| Clinical assessment (CCLAD & CDDP)** | | | 0.09 |
| TB assessment (CCLAD & CDDP)** | | | 0.09 |
| CDDP TB Case Finding Guide** | | | 0.04 |
| CDDP Drug Pick Up at community** | | | 0.04 |
| *Volunteer time only, not valued.  ** Personnel time for group events, split between all patients in the group. | | |  |
| **Laboratory test** | | **Per test** | |
| Viral Load | | 12.30 | |
| CD4 | | 9.60 | |
| TPHA (Syphilis test) | | 2.10 | |
| Complete Blood Count (CBC) | | 1.20 | |
| Malaria RDT | | 0.80 | |
| Serum Crag | | 4.10 | |
| GeneXpert | | 11.80 | |
| Haemoglobin | | 1.00 | |
| Urinalysis | | 0.80 | |
| HCG (Pregnancy test) | | 0.30 | |
| **First-line ARV regimens (fixed-dose combinations)** | **12 month supply***** | | |
| AZT/3TC/NVP | 73.61 | | |
| TDF/3TC/EFV | 76.04 | | |
| TDF/3TC/NVP | 77.26 | | |
| TDF/3TC/DTG | 99.16 | | |
| AZT/3TC/EFV | 100.38 | | |
| ABC/3TC/EFV | 114.37 | | |
| ABC/3TC/DTG | 124.71 | | |
| **Second-line regimens** | **12 month supply***** | | |
| ABC/3TC/NVP | 102.81 | | |
| AZT/3TC/DTG | 110.72 | | |
| TDF/3TC/ATV/r | 231.78 | | |
| AZT/3TC/ATV/r | 243.33 | | |
| ABC/3TC/ATV/r | 257.33 | | |
| TDF/3TC/LPV/r | 274.48 | | |
| AZT/3TC/LPV/r | 286.04 | | |
| ABC/3TC/LPV/r | 300.03 | | |

***Price includes supply chain management costs of 24.73%.

**S2 Table. Methods for cost estimation by cost category**

| Resource Input | Quantification method (Q) | Valuation technique (P) |
| --- | --- | --- |
| Variable costs (direct-patient resources reported in patient medical records and DSDM registers) | | |
| Medicines | Prescribed medicines during the study period, including ARVs and non ARVs. | The US$ (2018) unit price of medicines were used, as provided by the MOH Quantification Procurement & Purchasing Unit (QPPU). Total cost of drugs was estimated as quantity multiplied by unit cost. |
| Supply chain management costs | All medicines prescribed during the study period had a supply management cost applied to them. | The actual supply chain management cost varied depending on the medicine. An average mark-up of 24.725% on drug costs across all medicines was applied at the recommendation of the QPPU. |
| Viral load and other laboratory tests | Number of viral load (VL) tests and other laboratory tests done during the study period. | Unit price of VL test and others (US$2018) provided by the Central Public Health Laboratories (CPHL), and multiplied by the number of tests done over the period. |
| Human resources | The frequency of facility services or DSDM community events attributed to each patient were obtained from their ART care cards and the DSDM registers.  The different cadre engaged in providing specific types of facility services and DSDM events, and the average amount of time they each spent on these, were obtained through staff interviews – specific to each site and model-type. For group/community events, the average number of patients per group/ event were also obtained through staff interviews. | Public staff salaries were obtained from the public salary scales for 2017 and 2018. IP’s staff annual gross salaries (full cost to company) were obtained from their expenditure records. The cost per minute per cadre was calculated and applied to the reported amount of time (minutes) spent on facility services and DSDM community events. The HR cost per service/ event was multiplied by the numbers of each used by each patient. For group/ community events, the HR cost was divided by the average number of patients in the group/ event, and attributed to individual sampled patients.  A sensitivity analysis was performed to assess the possible range of under/over-reporting by staff. |
| Fixed costs (resources used for the facility and DSDM operation – both site-level and above-site) | | |
| Per diem and travel allowances for expert patient and staff | The number of per diems per model was based on the length of stay away from the facility, obtained through interviews with programme staff and volunteers. | The Government per diem rate (2018) and transport rate was applied to the number of support visits. |
| Vehicle maintenance costs | Through staff interviews the numbers of vehicles that were used in the DSDM service delivery were identified, and % of their maintenance costs that should be apportioned to DSDM activities was obtained. | Actual maintenance costs were obtained from the IP expenditure records, and the % attributed to the DSDM. By step down costing approach, these were then attributed to each patient in the DSDM. |
| Printing and record keeping | Total stationary, printing and files consumed in each year and their costs were based on actual expenditure. Through staff interviews the relevant % that should be apportioned to DSDM activities was obtained. | A proportion of the actual annual expenditure was applied for each specific model based on staff interviews, and then attributed to each patient in the DSDM (step down allocation). |
| Training costs | These included venue hire, per diem, travel costs, meals and refreshments and the development and supply of all training materials. Through staff interviews the relevant % that should be apportioned to specific sites and models. | Valuation was based on actual expenditure by the selected site or implementing partner (IP).  If training was a once-off cost before establishing the models, these costs were considered as capital investment and were discounted and annualized over a 3-year period. |
| Materials for repackaging of drugs | Actual quantities of the different materials used for repackaging of drugs were obtained through review of records. Through staff interviews the relevant % that should be apportioned to DSDM activities was obtained. | Quantities of different materials multiplied by their unit prices, obtained from the IP expenditure records. |
| Communication costs | Actual expenditure on communication that was relevant to DSDMs. | Where necessary, communication expenditure was apportioned to specific sites and models (if the expenditure was not separately kept for these models). |
| Overheads/  utilities | Actual expenditure on overheads for the site from expenditure records.  Through staff interviews the relevant % that should be apportioned to DSDM activities was obtained. | Apportioned total overheads expenditure to DSDM using the relevant %. |
| Facility and DSDM programme management (PM) & administration costs, incl. oversight and supervision | Actual expenditure on programme managers/ admin staff from expenditure records.  Through staff interviews the relevant % that should be apportioned to DSDM activities was obtained. | Apportioned total PM costs expenditure to DSDM using the relevant %.  A proportion of all these shared costs was then allocated to each DSDM patient using an allocation factor, per site and model, to obtain an average indirect cost per patient. This allocation factor was calculated as the numbers of ART visits per annum / total number of out-patient visits per annum at the facility. |
| Materials and supplies | Actual quantities of the different materials used for DSDM start up and for on-going operations. | Quantities of different materials multiplied by their unit prices (from IP expenditure records). |
| IEC materials | Actual expenditure on IEC materials for DSDMs, shared across DSDMs if more than one was being implemented | Quantities of different materials multiplied by their unit prices (from IP expenditure records). |
| Community sensitization and mobilization | Actual expenditure on mobilization costs per site from records. Through staff interviews the relevant % that should be apportioned to DSDM activities was obtained. | Apportioned total IP expenditure to DSDM using the relevant %. |
| Capital Costs | | |
| Vehicles / Motorcycles / Bicycles | Quantification of vehicles/motorcycles/bicycles were done through the KIIs, and their share of use for the specific models estimated (if not fully utilized by the model). | Current replacement costs for vehicles was obtained from MOH procurement price list. The estimated annual cost was annualized value of the vehicles. |
| Buildings | Space consumed by DSDM activities was measured in square meters. | A square meter was valued at UGX 61,240 (discounted and annualized), which was provided by the MOH (budget framework). |

##### S3 Table. DSDM patient’s ARV formulations (as at the end of 24-month study period)

| **ARV formulation** | **CCLAD (n=130)** | **CDDP (n=132)** | **FBG (Gp2) (n=115)** | **FBIM (n=126)** | **FDR (n=132)** | **Total (n=635)** |
| --- | --- | --- | --- | --- | --- | --- |
| **First-line regimens** | 95% | 92% | 99% | 90% | 84% | 92% |
| *AZT/3TC/NVP* | *28%* | *32%* | *12%* | *18%* | *21%* | *22.7%* |
| *TDF/3TC/EFV* | *35%* | *25%* | *82%* | *60%* | *32%* | *45.8%* |
| *TDF/3TC/NVP* | *13%* | *8%* | *4%* | *1%* | *12%* | *7.9%* |
| *TDF/3TC/DTG* | *12%* | *17%* | *1%* | *1%* | *8%* | *8.0%* |
| *AZT/3TC/EFV* | *5%* | *10%* | *0%* | *10%* | *11%* | *7.4%* |
| *AZT/3TC/DTG* | *1%* | *0%* | *0%* | *0%* | *0%* | *0.2%* |
| *ABC/3TC/EFV* | *1%* | *0%* | *0%* | *0%* | *0%* | *0.2%* |
| **Second-line regimens** | 5% | 8% | 1% | 10% | 16% | 8% |
| *TDF/3TC/ATV/r* | *1%* | *2%* | *1%* | *4%* | *3%* | *2.2%* |
| *AZT/3TC/ATV/r* | *0%* | *1%* | *0%* | *2%* | *2%* | *0.8%* |
| *ABC/3TC/ATV/r* | *2%* | *1%* | *0%* | *0%* | *6%* | *1.7%* |
| *TDF/3TC/LPV/r* | *2%* | *2%* | *0%* | *3%* | *3%* | *2.0%* |
| *ABC/3TC/LPV/r* | *0%* | *0%* | *0%* | *0%* | *2%* | *0.3%* |
| *AZT/3TC/LPV/r* | *1%* | *2%* | *0%* | *1%* | *1%* | *0.8%* |

**S4 Table. Types and frequency of diagnostic tests performed**

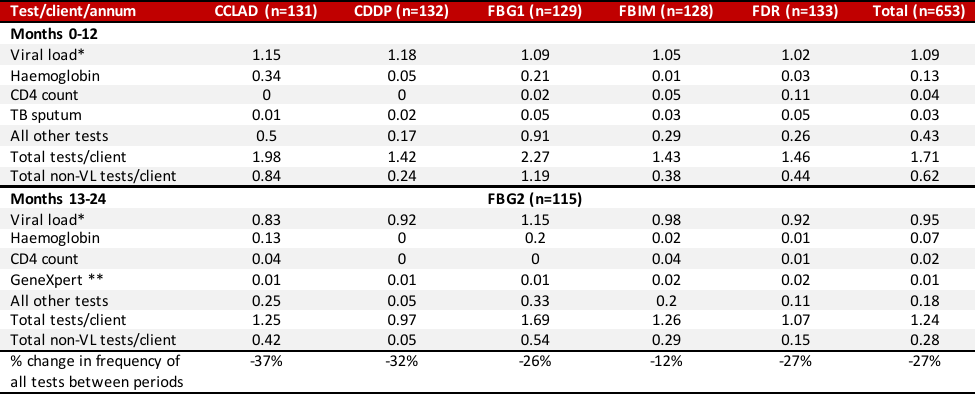

##### S5 Table. Types and frequency of facility-based services

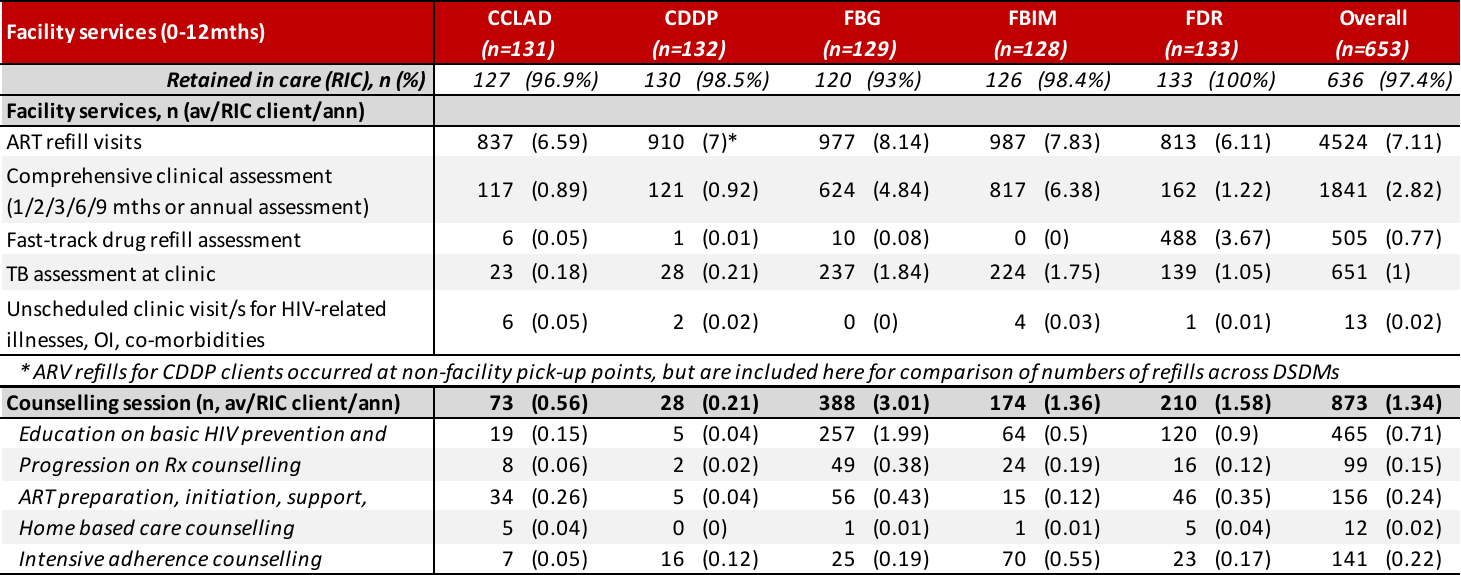

##### S6 Table. Types and frequency of DSDM events (non-facility based)

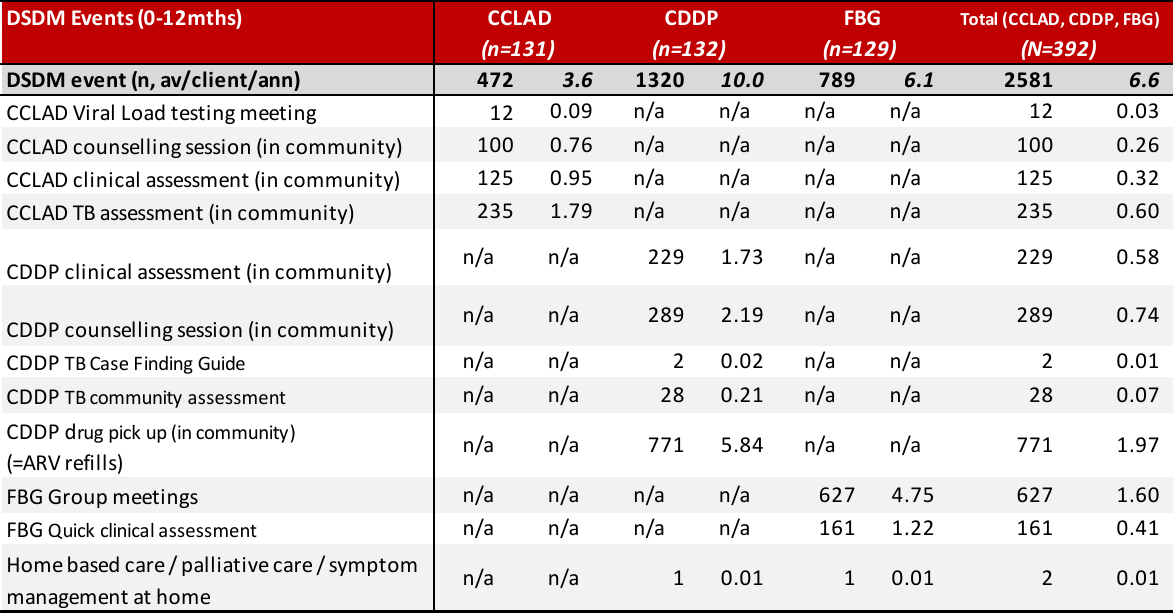

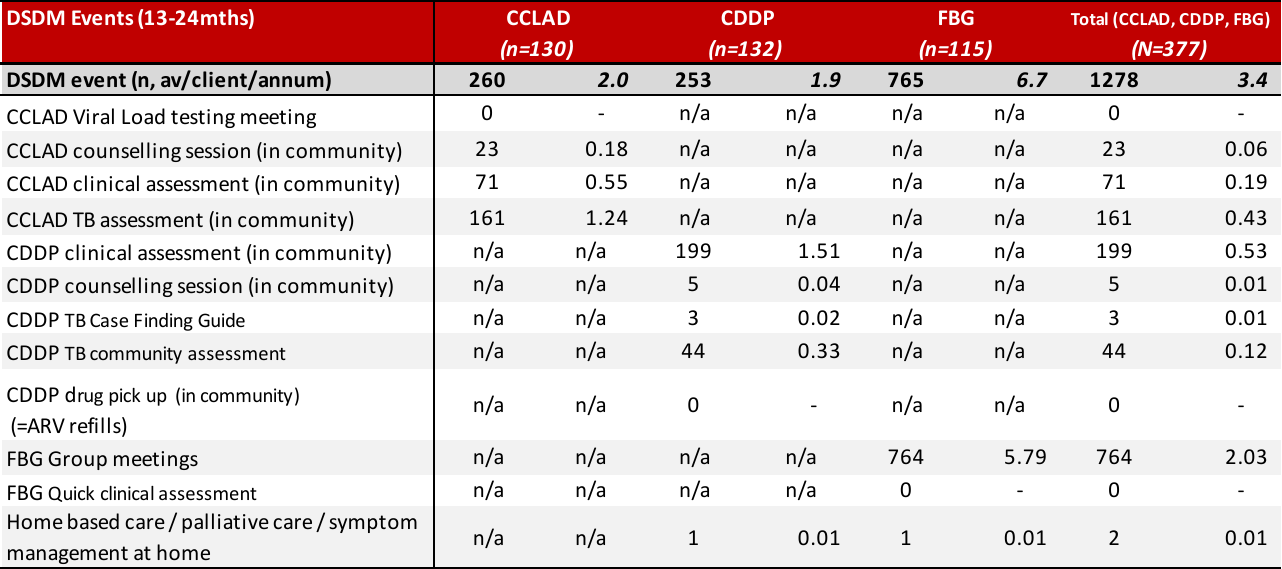

n/a = not applicable - where that particular model does not have those particular types of events.

**S1 File. Sampling: Selection of sites**

The study population for Phase 1 (top-down collection of implementers’ operational costs) were all the DSDMs in Uganda that would have been in operation longer than 6 months (by October 2017, the data collection point). There were 783 health facilities/sites that were implementing DSDMs in Uganda, serving a total number of 175,000 clients (according to IP reports).

Multi-stage purposive sampling was applied, since a randomized representative sample was beyond the time and resources available for this rapid study.

The DSDM site’s operating length of time was considered: only sites that had been in operation for more than 6 months were included in the sampling frame. In addition, the size of the DSDM sites, in terms of the numbers of clients served by each model, was considered. Those that were defined as too large or too small (dependent upon the model type) were excluded. Table 1 shows the different DSDMs’ average numbers of clients, and indicates the exclusion rules applied for each. Models that were outliers (i.e. very small or very large) were excluded, as determined below.

**Table 1. Definition of DSDM sizes (based on numbers of clients by type of DSDM) and exclusion criteria**

|  | CCLAD | CDDP | FBG | FDR |
| --- | --- | --- | --- | --- |
| Average # clients per group or site | 8 | 33 | 49 | 685 |
| *Rule for exclusion: min # of clients* | <4 | <10 | <10 | <100 |
| *Rule for exclusion: max # of clients* | >12 | >400 | >100 | >1000 |
| # sites excluded from sampling frame | 0 | 49 | 64 | 64 |

After these exclusions, 605 eligible sites remained in the sampling frame for inclusion. From these, 47 were selected through stratified purposive sampling.

The eligible models were first stratified by the type of model, and approximately 7%-8% from the CCLADs, CDDPs, FBGs and FDRs were selected. For the FBGs, additional effort was made to include their different client groups (pregnant and lactating women, children and adolescents, families). Only 4 FBIMs sites were added (at the request of the MOH), so as to include those clients who did not, or could not, join the other DSDMs, either by choice or by virtue of being complex (according to the Guidelines).

Within the model-types, the sites were then clustered according to their size (number of clients), and sites selected so as to have more or less similar amounts of each size, as shown in Table 2.

**Table 2. Defining site size by client numbers, and sites selected in each cluster**

| Sites’ number of clients | CCLAD | CDDP | FBG | FDR | Total in sample |
| --- | --- | --- | --- | --- | --- |
| Definition: Small | 4-6 | 10-30 | 10-20 | 100-400 | 12 |
| Definition: Medium | 7-9 | 31-80 | 21-50 | 401-700 | 19 |
| Definition: Large | 10-12 | 81-400 | 51-100 | 701-1000 | 12 |

* NB. The size of the 4 selected FBIM’s were not known at the time of sampling, and therefore the clustering by size did not apply to the FBIMs.

Sites were then purposively sampled based on their location, so as to ensure adequate amounts from each of the four Ugandan regions. The most extreme locations and hard to reach sites were omitted due to time and resource constraints. Finally, the selection of the 47 sites was made so as to include 10 different IPs, as shown in Table 3.

**Table 3. Number of sites per IP and per model included in the sample**

| Service Provider | CCLAD | CDDP | FBG | FDR | FBIM | Total |
| --- | --- | --- | --- | --- | --- | --- |
| ASSIST North | 2 |  |  |  |  | 2 |
| IDI |  | 1 | 4 | 1 | 6 | 12 |
| MILDMAY UGANDA | 2 | 1 | 1 |  |  | 4 |
| RHITES-EC | 1 |  |  |  |  | 1 |
| TASO | 2 | 1 | 3 | 1 |  | 7 |
| UEC/UCMB |  | 2 |  | 3 | 1 | 6 |
| UPMB |  | 1 | 1 |  |  | 2 |
| PIDC-COE-BAYLOR | 1 |  |  | 1 |  | 2 |
| RHITES-SW |  |  | 6 |  | 2 | 8 |
| REACH-OUT MBUYA |  | 1 |  | 1 | 1 | 3 |
| Total Ph1 sample sites | **8** | **7** | **15** | **7** | **10** | **47** |
| * Many of the Baylor-Uganda sites were excluded based on their short period of operation. Therefore, two of their 8 eligible sites were included.  IDI = Infectious Disease Institute]  UCMB = Uganda Catholic Medical Bureau  TASO = The AIDS Support Organization  UPMB = Uganda Protestant Medical Bureau  RHITES- EC = Regional Health Integration to Enhance Services- East and Central  RHITES- SW = Regional Health Integration to Enhance Services – South West | | | | | | |

In summary, Table 4 indicates the characteristics of the DSDM sites that were considered in the stratified purposive sampling. Refer to the following Table 5 below for the detailed names of sites and their locations.

**Table 4. Summary characteristics of sites**

| Site characteristic | Number included in sample | Percentage in sample |
| --- | --- | --- |
| DSDM type: |  |  |
| CCLAD | 9 | 19% |
| CDDP | 6 | 12.5% |
| FBG | 22 | 47% |
| FDR | 6 | 12.5% |
| FBIM | 4 | 9% |
| Regional location: |  |  |
| North | 9 | 19% |
| Central | 17 | 36% |
| East | 5* | 11% |
| West | 16 | 34% |
| Implementing Partner: | |  |
| ASSIST NORTH | 2 | 4.2% |
| BAYLOR | 2 | 4.2% |
| IDI | 12 | 25.5% |
| MILDMAY | 4 | 8.5% |
| REACHOUT MBUYA | 3 | 6.4% |
| RHITES-EC | 1 | 2.1% |
| RHITES-SW | 8 | 17% |
| TASO | 7 | 15% |
| UCMB | 6 | 12.8% |
| UPMB | 2 | 4.3% |
| Facility type: |  |  |
| Hospital | 12 | 26% |
| Health Centre IV | 11 | 23% |
| Health Centre III | 13 | 28% |
| TASO sites | 6 | 13% |
| Facility ownership: |  |  |
| Government | 26 | 55% |
| Private not for profit (PFNP) | 21 | 45% |
| * East region had fewer sample sites due to the large number of DSDMs operated there by TASO, who are conducting their own cost-efficiency analysis and therefore did not require as many sites included in this study. | | |

**Table 5. List of 47 sampled sites**

| Implementing Partner | Region | District | Health Facility | Health facility ownership | Type of DSDM |
| --- | --- | --- | --- | --- | --- |
| ASSIST NORTH | NORTH | LIRA | OGUR HC IV | GOVT | CCLAD |
| ASSIST NORTH | NORTH | OYAM | ANYEKE HC IV | GOVT | CCLAD |
| BAYLOR | CENTRAL | KAMPALA | BAYLOR HOSPITAL | PNFP | CCLAD |
| BAYLOR | CENTRAL | KAMPALA | BAYLOR HOSPITAL | PNFP | FDR |
| IDI | CENTRAL | KAMPALA | KISWA HC III | GOVT | CDDP |
| IDI | CENTRAL | MASINDI | KYATIRI HC III | GOVT | FBG |
| IDI | CENTRAL | KAMPALA | KOMAMBOGA HC III | GOVT | FBG |
| IDI | CENTRAL | KAMPALA | KISUGU HC III | GOVT | FBG |
| IDI | CENTRAL | KAMPALA | KISWA HC III | GOVT | FBG |
| IDI | CENTRAL | KIBOGA | LWAMATA HC III | GOVT | FBIM |
| IDI | WEST | HOIMA | BUHIMBA HC III | GOVT | FBIM |
| IDI | NORTH | ADJUMANI | ADJUMANI HOSPITAL | GOVT | FBIM |
| IDI | NORTH | ADJUMANI | DZAIPI HC III | GOVT | FBIM |
| IDI | WEST | MASINDI | BWIJANGA IV | GOVT | FBIM |
| IDI | CENTRAL | KAMPALA | KISWA HC III | GOVT | FBIM |
| IDI | CENTRAL | KAMPALA | IDI MULAGO | PNFP | FDR |
| MILDMAY | CENTRAL | MITYANA | MITYANA GENERAL HOSPITAL | GOVT | CCLAD |
| MILDMAY | CENTRAL | WAKISO | MILD MAY HOSPITAL | PNFP | CCLAD |
| MILDMAY | CENTRAL | WAKISO | KAJJANSI HC IV | GOVT | CDDP |
| MILDMAY | CENTRAL | WAKISO | KIRA HC III | GOVT | FBG |
| REACHOUT MBUYA | CENTRAL | KAMPALA | REACHOUT MBUYA | PNFP | CDDP |
| REACHOUT MBUYA | CENTRAL | LUWERO | St. MARY'S KASALA | PNFP | FBIM |
| REACHOUT MBUYA | CENTRAL | LUWERO | St. MARY'S KASALA | PNFP | FDR |
| RHITES -EC | EAST | MAYUGE | KITYERERA HC IV | GOVT | CCLAD |
| RHITES -SW | WEST | IBANDA | NYAMAREBE HC III | GOVT | FBG |
| RHITES -SW | WEST | BUSHENYI | BUSHENYI HC IV | GOVT | FBG |
| RHITES -SW | WEST | IBANDA | RUHOKO HC IV | GOVT | FBG |
| RHITES -SW | WEST | BUHWEJU | NSIIKA HC IV | GOVT | FBG |
| RHITES -SW | WEST | MITTOMA | MITOOMA HC IV | GOVT | FBG |
| RHITES -SW | WEST | SHEEMA | KITAGATA HOSPITAL | GOVT | FBG |
| RHITES -SW | WEST | ISINGIRO | NYARUBUNGO HC III | GOVT | FBG |
| RHITES -SW | WEST | IBANDA | NYAMAREBE HC III | GOVT | FBIM |
| RHITES -SW | WEST | BUSHENYI | BUSHENYI HC IV | GOVT | FBIM |
| TASO | WEST | MASINDI | TASO | PNFP | CCLAD |
| TASO | EAST | JINJA | TASO | PNFP | CCLAD |
| TASO | NORTH | GULU | TASO | PNFP | CDDP |
| TASO | EAST | TORORO | TASO | PNFP | FBG |
| TASO | EAST | JINJA | TASO | PNFP | FBG |
| TASO | EAST | MBALE | TASO | PNFP | FDR |
| UCMB | WEST | BUSHENYI | St. DANIEL COMBONI HOSPITAL | PNFP | CDDP |
| UCMB | NORTH | OYAM | POPE JOHN HOSPITAL ABER | PNFP | CDDP |
| UCMB | NORTH | OYAM | POPE JOHN HOSPITAL ABER | PNFP | FBIM |
| UCMB | WEST | BUSHENYI | St DANIEL COMBONI HOSPITAL | PNFP | FDR |
| UCMB | NORTH | OYAM | POPE JOHN HOSPITAL ABER | PNFP | FDR |
| UCMB | NORTH | GULU | LACOR HOSPITAL | PNFP | FDR |
| UPMB | WEST | HOIMA | AZUR HC IV | PNFP | CDDP |
| UPMB | WEST | MBARARA | RUHARO MISSIONARY HOSPITAL | PNFP | FBG |

* IDI = Infectious Disease Institute]

* UCMB = Uganda Catholic Medical Bureau

* TASO = The AIDS Support Organization

* UPMB = Uganda Protestant Medical Bureau

* RHITES- EC = Regional Health Integration to Enhance Services- East and Central

* RHITES- SW = Regional Health Integration to Enhance Services – South West

PFNP = Private not for profit

HC= Health Centre

### Sampling of the 20 sub-set: selection of sites

From the 47 sites, a sub-set of twenty sites were purposively selected for the collection of direct-patient resource utilization. This was primarily due to resource constraints which prohibited all 47 sites from being included. Four sites for each of the five types of DSDMs, i.e. CCLAD, CDDP, FDR, FBG, and FBIM were included, all of which had been in operation for at least a year by the time of the first study period (months 1-12) data collection and were being implemented by seven IPs.

**Table 6. List of 20 Sampled sites for Phase 2**

| Implementing partner | Number of sites of each model included | | | | | Total |
| --- | --- | --- | --- | --- | --- | --- |
|  | **CCLAD** | **CDDP** | **FBG** | **FBIM** | **FDR** |  |
| PIDC-COE-Baylor |  |  |  |  | 1 | 1 |
| IDI |  | 1 |  | 1 |  | 2 |
| MildMay |  |  | 1 | 1 |  | 2 |
| Reach Out Mbuya |  |  |  | 1 |  | 1 |
| RHITES – SW |  |  | 2 |  |  | 2 |
| TASO | 4 | 2 | 1 |  | 1 | 8 |
| UCMB |  | 1 |  | 1 | 2 | 4 |
| Total | **4** | **4** | **4** | **4** | **4** | **20** |
